# An automatic entropy method to efficiently mask histology whole-slide images

**DOI:** 10.1101/2022.09.01.22279487

**Authors:** Yipei Song, Francesco Cisternino, Joost M. Mekke, Gert J. de Borst, Dominique P.V. de Kleijn, Gerard Pasterkamp, Aryan Vink, Craig A. Glastonbury, Sander W. van der Laan, Clint L. Miller

## Abstract

**Background:** Tissue segmentation of histology whole-slide images (WSI) remains a critical task in automated digital pathology workflows for both accurate disease diagnosis and deep phenotyping for research purposes. This is especially challenging when the tissue structure of biospecimens is relatively porous and heterogeneous, such as for atherosclerotic plaques.

**Methods:** In this study, we developed a unique approach called `EntropyMasker` based on image entropy to tackle the fore- and background segmentation (masking) task in histology WSI. We evaluated our method on 97 high-resolution WSI of human carotid atherosclerotic plaques in the Athero-Express Biobank Study, constituting hematoxylin and eosin (H&E) and 8 other staining types.

**Results and Conclusion:** Using multiple benchmarking metrics, we compared our method with four widely used segmentation methods: Otsu’s method, Adaptive mean, Adaptive Gaussian and `slideMask` and observed that our method had the highest sensitivity and Jaccard similarity index. We envision `EntropyMasker` to fill an important gap in WSI preprocessing and deep learning image analysis pipelines and enable disease phenotyping beyond the field of atherosclerosis.

## Introduction

Atherosclerosis is a chronic inflammatory process resulting in arterial stiffening and plaque formation, and is the leading cause of myocardial infarction, ischemic stroke, and peripheral artery disease^1,2^. Historically, researchers and pathologists have characterized atherosclerotic plaque through standard histology and light microscopy analysis^3–6^. The composition of atherosclerotic plaques is highly variable, with different plaque types having distinct clinical manifestations^3,7^. For instance, more stable, fibrous-rich atheroma plaques are typically asymptomatic, whereas unstable, thin-cap fibroatheroma plaques are more prone to rupture and thrombus formation underlying cerebral or coronary events^5,6^. However, the value of atherosclerotic plaque composition in predicting cardiovascular events remains a subject of debate and ongoing research^7–9^.

While histology-based assessment of cancer and other diseases is often required for clinical diagnosis, histological analysis of atherosclerosis can reveal the extent of disease progression as well as underlying etiology. For example, in pre-clinical models of atherosclerosis, the amount of smooth muscle content and collagen extracellular matrix in the plaque often correlates with greater plaque stability^8^. These features can now be easily captured from stained tissue sections and digitized as high-resolution Whole Slide Images (WSI). WSI data provides a rich resource for quantitative and qualitative image analysis and has been a focus of digital pathology^10^. Overall, this has simplified archiving, enabled remote diagnosis, and accelerated both clinical decision-making and research investigations^11^.

Application of quantitative WSI analysis pipelines requires access to uniformly processed and well-characterized tissue biobanks. The Athero-Express study^3^ is a large-scale vascular tissue biobank comprising over 3,600 carotid and femoral endarterectomy surgical specimens, which include detailed clinical outcomes and follow-up. This has led to several prospective studies to correlate local atherosclerotic plaque composition with future local and systemic vascular outcomes, using histology^7–10^, RNA^12,13^, genetics^14^, and protein^15,16^ data. In the histology-based analyses, local plaque indices such as plaque hemorrhage and neovascularity were shown to correlate with more adverse vascular outcomes^7,9^.

Accurate tissue segmentation is a necessary initial step in WSI analysis in digital pathology workflows, particularly for deep learning-based classification. However, the process of WSI acquisition has its own technical challenges. Whole-slide scanners are known to fail to identify or “mask” the entire tissue region, e.g., due to dust specs, transparent tissue types (e.g., adipose) or weak immunostaining results. Omitted tissue areas could pose a significant problem for diagnostic accuracy, e.g., such as in cancer, identifying malignant regions in lymph nodes^17^. These tissue recognition mistakes may have costly downstream consequences in automated digital pathology workflows, be it for diagnostic or research purposes. By highlighting the tissue-specific area, deep learning algorithms can run more efficiently by only analyzing the informative portions of the WSI. For example, only 22 percent of the total WSI area was estimated in the CAMELYON17 challenge dataset^18^. In addition, inter- and intra-image color fluctuations, staining and slide preparation aberrations must also be considered through a robust tissue segmentation method to improve WSI interpretation.

### Masking in histological sections

The most straightforward method for identifying tissue on a white background is to set a specified threshold for the grayscale version of the image^19^, specific color channels^20^, or the optical density of the red, green, and blue (RGB) color channels^21^. Alternatively, WSIs can be split into a uniform image patch grid, then grayscale image thresholded, and the resultant binary tissue map used to reject image patches with insufficient tissue ^22,23^.

The most commonly used tissue masking method is Otsu’s adaptive thresholding ^24^, a technique applied to WSI based algorithm training in the CAMELYON17 challenge and other studies^18,25^. Otsu’s method has been extended by capturing additional thresholding areas from the edge of the binary mask^26^. In addition, several methods have been developed to improve WSI based tissue recognition. For instance, GrabCut^27^ for image segmentation of regions of interest, Watershed for cell or nuclei segmentation,^28^ and foreground extraction from structural information (FESI)^29^ for fore-and background segmentation. Thresholding at a fixed value and Otsu’s approach are easy to implement but have been difficult to generalize to more variable stains or clinical specimens. A more accurate approach is still needed in digital pathology workflows, especially for specimens containing small empty spaces between extracellular matrix fibers (e.g., collagen) or due to decalcifying procedures, and transparent tissue for example in adipose tissue or fat deposits in atherosclerotic plaques.

In this paper, we propose a fully automated approach for separating foreground (tissue) and background in histopathological bright-field microscopy images of hematoxylin and eosin (H&E)-stained samples. Our method, `EntropyMasker`, is unaffected by changes in scanning or image processing conditions, by using a measure of local entropy and generating corresponding binary tissue masks.

## Methods

### Our method: EntropyMasker

For uniform image processing, ideally the slide background is evenly achromatic. Unfortunately, in practice, there is often inconsistency of the background color due to variations in image capturing color temperature (e.g., from shades of blue to yellow), resulting in edge artifacts from tile-based scanning methods^30^. We corrected this background-color inconsistency by applying color hysteresis thresholding based on color distance from the “most common” background color. By doing so, we were able to easily determine the “seed regions” of the slide background (i.e., color range unique to the background).

In information theory^31^, entropy is defined as the log-base-2 of the number of possible outcomes for a sent/received message. In image analysis, entropy refers to the degree of randomness or complexity of pixels in a defined region or neighborhood of pixels. We defined the local entropy of a specific region as follows:

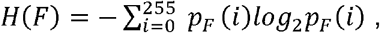

where *p*_*F*_ (*i*) represents the probability of a grayscale pixel, *i*, in a local footprint region.

In the case of *p*_*F*_ (*i*) = 0 for some *i*, the corresponding sum value and 0*log*_2_0 is set to *0*, which is consistent with the limit: 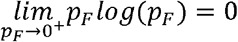.

Based on this approach, informative pixels have a low probability and contribute less to the local entropy, whereas noise pixels have a high probability and contribute more to the local entropy. Also, the background of an image has lower entropy due to higher brightness and more homogeneous texture, whereas foreground tissue has higher entropy due to the diversity of pixel intensities in the local neighborhood.

To extract tissue from the background of the image, we applied a filter to the generated local entropy value map. The filter is a threshold for determining the optimal range of local entropy values to distinguish background noise from tissue in the WSI. Since the background of the image has a lower entropy value, we set the threshold as the local minima of the histogram of the entropy map. We then assigned a Boolean value to each pixel in the WSI, defined as background or not, based on the entropy values compared to the threshold. These binary values were used to create the tissue segmentation mask for each WSI.

### Otsu’s method

Otsu’s method is a clustering-based image thresholding algorithm^24^, commonly utilized in image analysis and digital histopathology applications^25,30,32,33,34^. Following a bimodal histogram, the technique assumes that the image has two classes of pixels (tissue pixels and background pixels). It determines the best threshold for dividing the two groups such that their total intra-class variance is as low as possible.

### Adaptive thresholding

Adaptive thresholding is another widely used method for calculating the threshold value for smaller areas of WSIs^35,36,37,38^. Typically, these threshold values are calculated in two different ways, either using the mean of the neighborhood area, or a Gaussian weighted sum of neighborhood values.

### slideMask method from slideToolKit

The slideToolKit^39^ is an assistive tool set for the histological quantification of whole-slide images (https://github.com/swvanderlaan/slideToolKit). The `slideMask` tool in slideToolKit uses `convert` from ImageMagick (https://imagemagick.org) and a miniature version of the WSI to generate a mask (https://github.com/swvanderlaan/slideToolKit/blob/master/slideMask). The image is first blurred to eliminate dust and speckles. Then `Fuzz` in ImageMagick^40^ (which is computed as the root mean squared difference between two colors of the pixels) is used to create a fuzzy, non-stringent selection to identify the white background, which is subsequently substituted with black. The `slideMask` tool has settings for blur and fuzziness that may be explored and altered. Manual adjustments may be made to generated masks in any image editor (such as free GNU Image Manipulation Program; GIMP^38,41^). Unwanted regions on the WSI (such as marker stripes or air bubbles beneath the coverslip) may necessitate this procedure.

All proposed methods were implemented in the Python programming language using OpenCV^42^ and other required packages (see **Data and code availability** for more details).

### Athero-Express Biobank Study

#### Patient population

Atherosclerotic plaques were obtained from patients undergoing a arterial endarterectomy procedure and included in the Athero-Express Biobank Study (AE), an ongoing biobank study at the University Medical Centre Utrecht (Utrecht, The Netherlands) and the St. Antonius Hospital (Nieuwegein, The Netherlands)^3^. The medical ethical committees of the respective hospitals approved the AE which was registered under number TME/C-01.18. This study complies with the Declaration of Helsinki, and all participants provided informed consent. However, considering national laws plaque-material are considered ‘waste biomaterial’ and are always allowed to be used without any personal information regardless of informed consent. In this study we also considered this ‘waste biomaterial’, hence no clinical information, e.g., age, is given here nor when sharing data of the relevant samples (n=3). The study design of the AE was described before^3^, but in brief: during surgery blood and plaques are obtained, stored at -80°C and plaque material is routinely used for histological analysis^3,43^.

#### Whole-slide staining and scanning

The standardized (immuno)histochemical analysis protocols used in the AE biobank have been described previously.^3,43^ In short, 4-micron cross sections of the paraffin-embedded segments were cut using a microtome, and 8 different stains were applied for endothelial cells (CD34), macrophages (CD68), elastic Van Gieson (EvG), fibrin, red blood cells (glycophorin C, GLYCC), hematoxylin and eosin (H&E) for nuclei, collagen (picrosirius red, SR), and smooth muscle cells (SMCs, α--actin) on consecutive slides. We set up ExpressScan to obtain whole-slide images (WSIs) by scanning stained slides at 40x using a Roche Ventana iScan HT (https://diagnostics.roche.com/global/en/products/instruments/ventana-iscan-ht.html) or Hamamatsu C12000-22 Digital slide scanner (https://www.hamamatsu.com/eu/en/product/life-science-and-medical-systems/digital-slide-scanner/index.html). WSIs were stored digitally as z-stacked `.TIF` (Roche) at 0.25 micron/pixel or `.ndpi` (Hamamatsu) at 0.23 micron/pixel brightfield microscopy images^10^. The slides numbers used for this project are listed in **Supplemental Table 1**.

### Manual tissue annotation and masking

The ground truth tissue areas for each WSI were annotated manually at 40x magnification using QuPath^44^. If the tissue had disintegrated into several disjointed fragments during preparation or staining the annotators marked them with a single enclosing polygon. Binary masks of these tissue annotations were then generated using groovy scripts in QuPath.

### Statistical analyses

We assessed the algorithms using several metrics: The Jaccard index, sensitivity, False positive Rate (FPR) and pixel accuracy, comparing the output of the algorithms to the ground truth binary reference masks of the 97 images in the test set.

Jaccard index was applied to assess the performance of our masking algorithms, by measuring the overlap area of target masks with the ground truth masks annotated by experts and masks of the algorithm outputs divided by the area of the union of both masks. Mathematically, sensitivity, false positive rate and pixel accuracy can be expressed as:

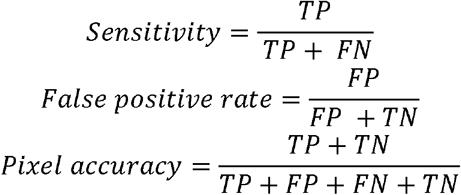

### Data and code availability

The histological data as used here are available through https://doi.org/10.34894/GN4YOS.

All documented code and tutorial to run `EntropyMasker` can be found here https://github.com/CirculatoryHealth/EntropyMasker.

### Ethics statement

The studies involving human participants were reviewed and approved by Medisch Ethische Toetsingscommissie (METC) Utrecht. The patients/participants provided their written informed consent to participate in this study.

### Evaluation and results

We developed `EntropyMasker`, a novel masking method using entropy-filtering (**Figure 1**). First, we compared our method against three conventional thresholding methods: Otsu’s, adaptive thresholding using the mean of neighborhood area as the threshold, and adaptive thresholding using the Gaussian weighted sum of neighborhood values as the threshold. We also evaluated these methods against the previously developed `slideMask` tissue masking method in the slideToolKit^39^.

**Figure 1:**
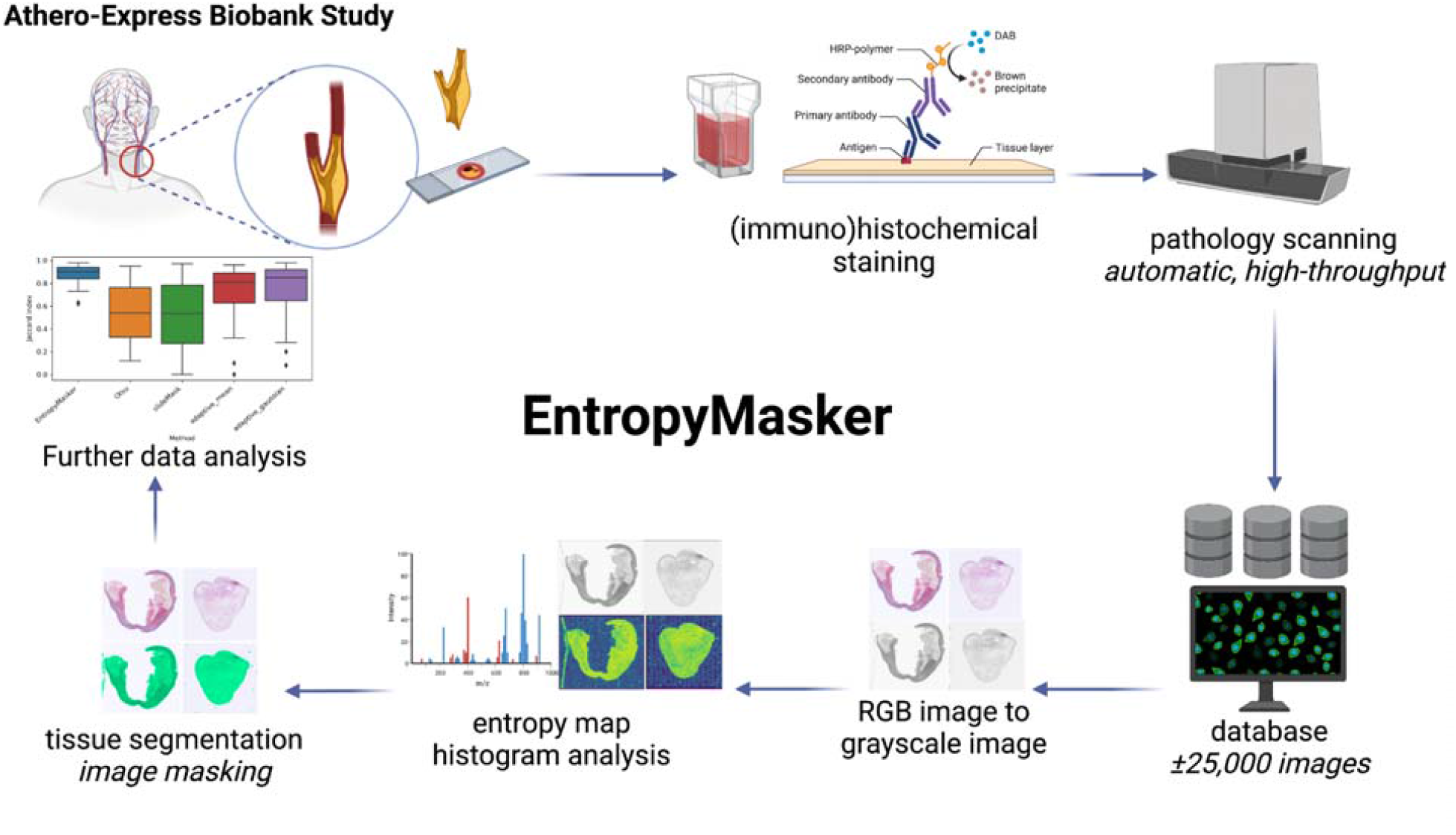
Schematic overview of the EntropyMasker method for masking whole-slide images. Atherosclerotic plaque tissue samples were collected from arterial endarterectomy surgeries, sectioned, and mounted on tissue slides as part of the Athero-Express Biobank Study. The slides were subjected to histological staining for overall morphology (hematoxylin & eosin; H&E), collagen deposition (Picrosirius red; SR), fibrin deposition (Fibrin), elastin fibers (Elastica Van Gieson; EVG) or immunohistochemical staining for cell-type specific markers: alpha-smooth muscle actin (a-SMA), macrophages (CD68), endothelial cells (CD34), or erythrocytes (glycophorin C). Whole-slide images of each slide were generated using high-resolution scanners and images were stored in a database (∼25,000 images). Red, Green, Blue (RGB) color images were converted to grayscale images and grayscale pixels were used to calculate entropy threshold map for histogram analysis, and tissue segmentation was performed to create tissue mask of WSIs for subsequent data analysis and benchmarking. Schematic was created with BioRender.com

We evaluated each method using 97 WSIs (**Supplemental Table 1** for details on usage and **Supplemental Table 2** for a minimalistic baseline table) at 40X magnification on layer 6 (pixel spacing at layer 6 is around 8.36µm) consisting of 8 different stains: H&E, picrosirius red, Fibrin, EVG, a-SMA, CD34, CD68, and GLYCC. We did not apply our method to layer 1 due to memory constraints. The average Jaccard index, also referred to as the Intersection over Union (IoU) metric, of the adaptive thresholding method where its threshold value is the weighted sum of neighborhood values where weights are a gaussian window was 0.7702 and the average sensitivity was 0.8597. This masking method produced the highest pixel accuracy among all of the traditional methods which was 0.9485. Otsu’s method had a much lower false positive rate than any of the approaches examined at the cost of much lower average Dice scores and sensitivity levels. The quality of the outcome of the `slideMask` algorithm was unevenly distributed resulting in a relatively low Jaccard index (IoU) across all the tested methods. The average false positive rate was 0.0153, which is the lowest of all masking methods in comparison, and its average pixel accuracy was 0.9133.

Our automatic local entropy-based masking method, `EntropyMasker`, had the best average Jaccard index (IoU) 0.8837 and the highest sensitivity 0.9685, outperforming Otsu’s method, two adaptive methods and `slideMask` masking method in both metrics. The average false positive rate was 0.0176 which is much lower than Otsu’s method, and two adaptive methods. **Table 1** contains the complete set of metric values and **Figure 2** shows the summary statistics of the Jaccard index across all 97 slides. The results of comparing the 5 masking methods for an H&E-stained whole-slide image are shown in **Figure 3** where generated tissue masks are overlaid by pseudocolor (green).

**Table 1:** Comparison of different masking methods. The method with the highest evaluation metric is shown in bold. Metric values represent mean ± SD.

| Method | Jaccard index | Sensitivity | False positive rate | Pixel accuracy |
| --- | --- | --- | --- | --- |
| `slideMask` | 0.5273±0.29 | 0.5451±0.30 | 0.0153±0.06 | 0.9133±0.08 |
| Otsu's | 0.5499±0.25 | 0.5608±0.25 | 0.021±0.09 | 0.8994±0.10 |
| Adaptive_mean | 0.7322±0.21 | 0.7691±0.24 | 0.0278±0.06 | 0.9452±0.06 |
| Adaptive_gaussian | 0.7702±0.20 | 0.8597±0.18 | 0.0393±0.09 | 0.9485±0.07 |
| **EntropyMasker** | **0.8837±0.08** | **0.9685±0.06** | **0.0176±0.02** | **0.9111±0.07** |

**Figure 2:**
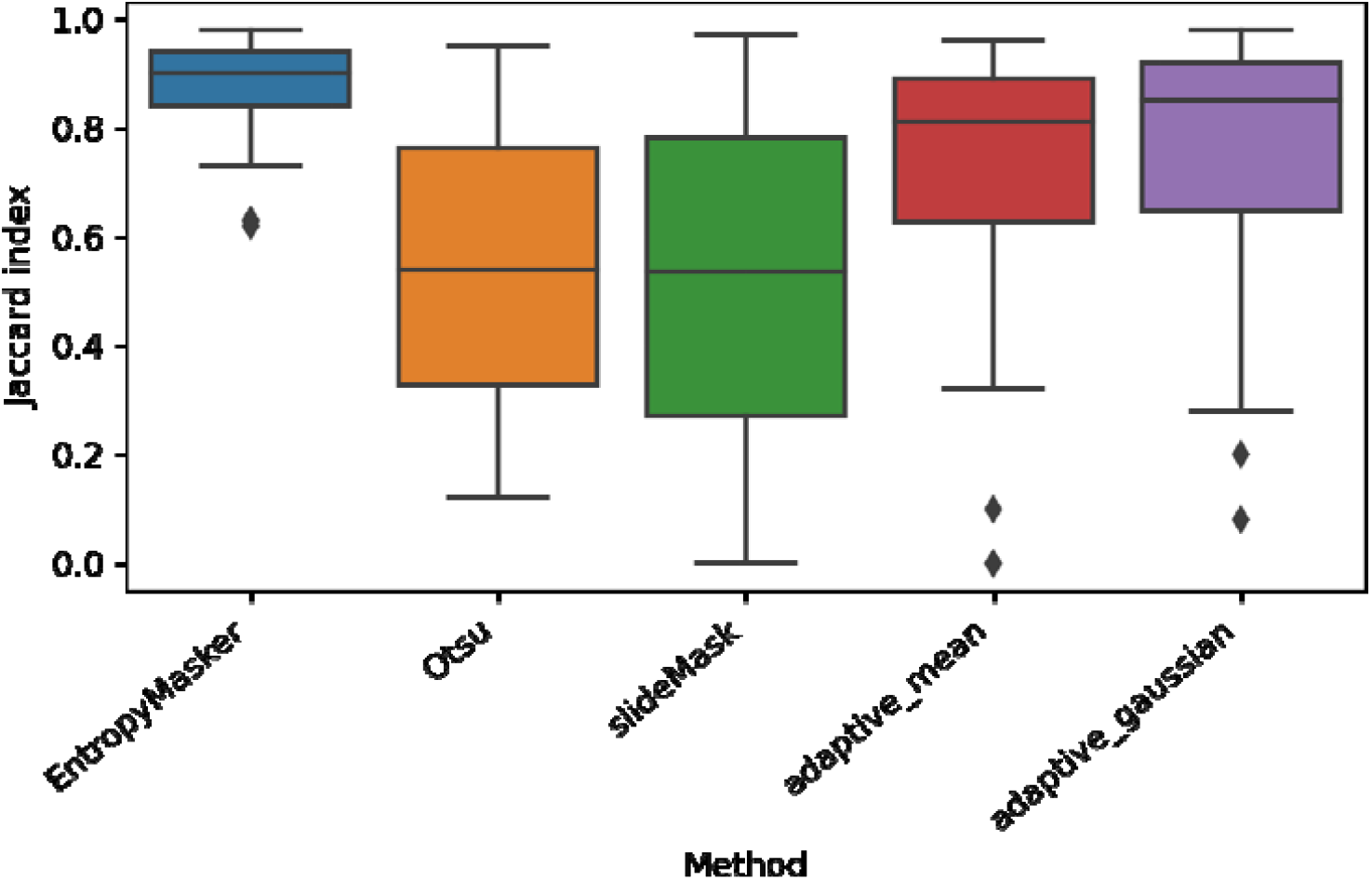
Jaccard index for the different methods used. Comparison of `EntropyMasker` with other masking methods, Otsu, `slideMask`, and adaptive thresholding methods, using manually annotated tissue sections as ground truth and calculated Jaccard index (intersection over union metric). 97 images were used from 65 individual patient samples to evaluate each method. Boxes represent the median and interquartile range (IQR) and whiskers represent up to 1.5X IQR. Outliers (greater than 1.5X IQR) are shown as diamonds.

**Figure 3:**
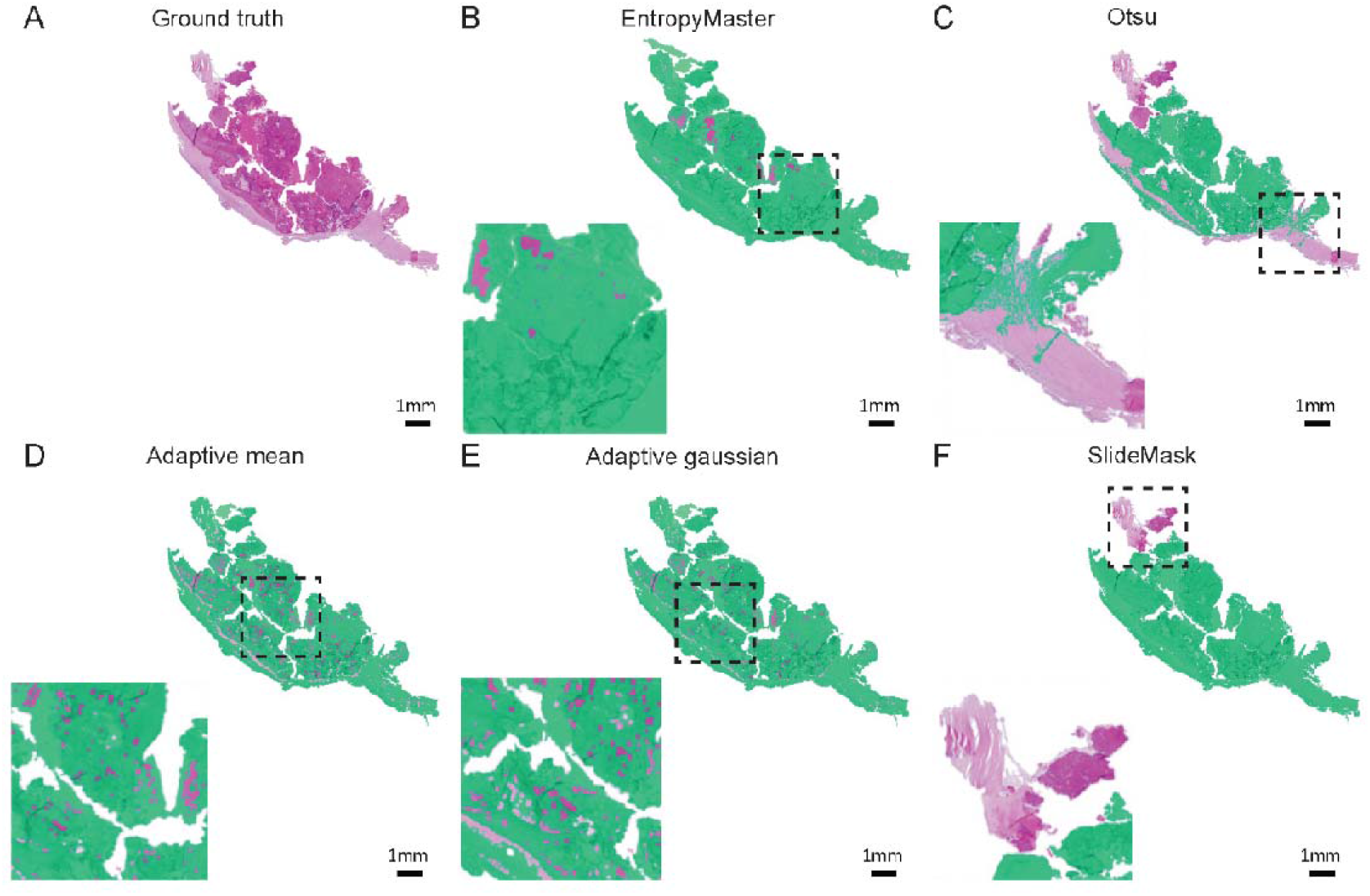
Comparison of results of 5 masking methods for an H&E-stained whole-slide image. A) Expert generated annotation (ground truth) using Qupath Software (Version 0.3.2)^44^. B) Our `EntropyMasker` method. C) Otsu’s method. D) Adaptive method where the threshold value is the mean of neighborhood area. E) Adaptive method where the threshold value is the weighted sum of neighborhood values where weights are a gaussian window. F) `slideMask` method. All the generated tissue masks are overlaid by pseudocolor (green).

To demonstrate the generalizability of our automatic entropy method, we tested on whole-slide images with 8 different types of staining. Examples of masking results of our proposed method are shown on **Figure 4** where generated tissue masks are overlaid by pseudocolor (green).

**Figure 4:**
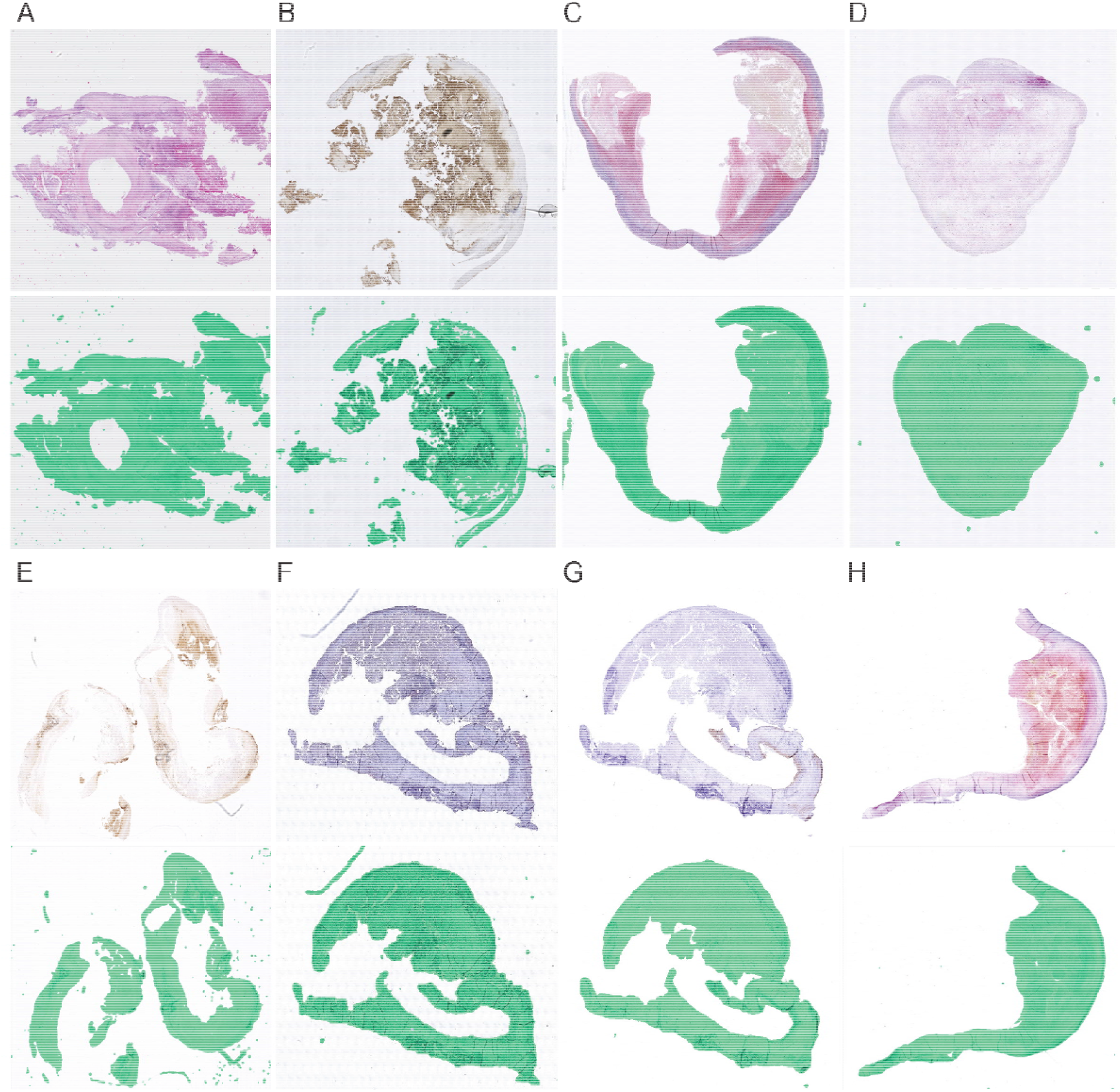
Generalizability with different antibody specific stained sections. Images of the upper part are the original whole slide images at layer 6 with 8 different types of staining methods respectively and images of the lower part are the examples of masking results of our `EntropyMasker` method where generated tissue masks are overlaid with pseudocolor (green). (A) H&E; (B) Glycophorin C; (C) Sirius red; (D) Fibrin; (E) CD68; (F) CD34; (G) Smooth muscle actin; (H) EVG.

The computation speed for Otsu’s method, adaptive methods, `slideMask` and proposed `EntropyMasker` method on a 3.6GHz Quad-Core Intel Core i7 processor, 32GB 2400MHz DDR4 memory iMac are 0.08, 0.11, 0.25 and 0.34 seconds per one million pixels, respectively.

## Conclusions and discussion

Overall, our automatic entropy masking method, `EntropyMasker`, performed well on atherosclerotic plaque cross-sectional WSIs derived from 8 different types of stains. With a relatively high average Jaccard index, our method was able to separate out the foreground and background accurately and consistently from these complex images (**Figure 2 and 3**).

In comparison, we observed the popular masking method, Otsu’s, and our previous method, `slideMask`, both tended to miss some tissue areas along the tissue boundary. This likely results from the relatively lower intensity of these areas, where Otsu’s method sets a threshold in the middle of two peaks, thus partially omitting the higher intensity pixels compared to the threshold (**Figure 3**). The `slideMask` method computes the root mean squared difference between two colors, however this becomes problematic when the intensity of tissue area pixels is relatively large (**Figure 3**).

We also found that the adaptive methods tend to produce ‘porosity’ during the masking, since these methods miss small regions of the image which are supposed to be regarded as tissue (**Figure 3**). These large-scale missing tissue makes Otsu’s, `slideMask` and adaptive thresholding methods impractical for use in many image processing workflows^10,39^, especially when applied to atherosclerotic plaque images. Given the limited availability and heterogeneous nature of these plaques, an ideal masking method will retain all the plaque components, which can be used to discover tissue or compartment-specific markers for disease progression and phenotyping. This is also a valuable feature of `EntropyMasker`, which can be implemented at the preprocessing stage of various deep learning pipelines for other diseases and research areas which require a complete WSI masking step^45–47^.

Despite the accurate and consistent performance of `EntropyMasker`, it is worth pointing out some known limitations. In comparison to the other methods that we evaluated, our method has the highest sensitivity, and thus tends to over-include a few pixels outside the edges of tissue (**Figure 4**). While these regions represent a small fraction of the total tissue areas that we tested, this should be more carefully considered when evaluating smaller tissue sections^11^. Also, our method had slightly lower pixel accuracy than the adaptive methods, which may be important for tasks involving more subtle intensity differences in defining target regions of the tissue^48^. Compared to the other methods discussed, ours is slightly slower (0.34 seconds per one million pixels compared to 0.11 seconds for Otsu’s) which may pose an issue when examining thousands of images, although this should be negligible since our method is scalable on any high-performance compute cluster.

In conclusion, we demonstrated the effectiveness of our proposed method for tissue masking on human atherosclerotic plaque WSIs of different types of stains including H&E, CD34, CD68, EVG, smooth muscle cell α-actin, picrosirius red, fibrin and Glycophorin C. By evaluating our method against other popular masking methods and those recently developed, we also demonstrated that our entropy-based masking method is scalable and had optimal performance across these WSIs. Thus, we envision `EntropyMasker` to serve an important role in WSI preprocessing and deep learning image analysis pipelines and enable disease phenotyping beyond the field of atherosclerosis.

## Data Availability

The histological data as used here are available through https://doi.org/10.34894/GN4YOS.
All documented code and tutorial to run EntropyMasker can be found here https://github.com/CirculatoryHealth/EntropyMasker.

## Author contributions

We used the CRediT taxonomy (https://journals.plos.org/plosgenetics/s/authorship#loc-author-contributions) to determine the author’s contributions summarized below.

| Contributor Role | YS | FC | JMM | GJd<br>B | DPVdK | GP | AV | CAG | SWvd<br>L | CLM |
| --- | --- | --- | --- | --- | --- | --- | --- | --- | --- | --- |
| **Conceptualization** | x |  | x |  |  |  |  |  | x | x |
| **Data Curation** | x |  | x |  |  |  |  | x | x |  |
| **Formal Analysis** | x |  |  |  |  |  |  |  |  |  |
| **Funding Acquisition** |  |  |  |  |  |  |  |  | x | x |
| **Investigation** | x |  |  |  |  |  |  |  |  |  |
| **Methodology** |  |  |  |  |  |  |  |  |  |  |
| **Project Administration** |  |  |  |  |  |  |  | x | x | x |
| **Resources** |  |  | x | x | x | x | x |  | x |  |
| **Software** | x |  |  |  |  |  |  | x | x |  |
| **Supervision** |  |  |  | x | x | x |  | x | x | x |
| **Validation** |  | x |  |  |  |  |  |  |  |  |
| **Visualization** | x |  |  |  |  |  |  |  | x | x |
| **Writing – Original Draft Preparation** | x |  |  |  |  |  |  |  |  |  |
| **Writing – Review & Editing** | x | x | x | x | x | x | x | x | x | x |

## Acknowledgements

We also acknowledge support from the Netherlands CardioVascular Research Initiative of the Netherlands Heart Foundation (CVON 2011/B019 and CVON 2017-20: Generating the best evidence-based pharmaceutical targets for atherosclerosis [GENIUS I&II]), the ERA-CVD program ‘druggable-MI-targets’ (grant no: 01KL1802), and the Chan Zuckerberg Initiative Foundation Data Insights program (grant no: DI-092).

We would also like to thank all the (former) employees involved in the Athero-Express Biobank Study of the Departments of Surgery of the St. Antonius Hospital Nieuwegein and University Medical Center Utrecht for their continuing work. We would like to thank (in no particular order) Marijke Linschoten, Arjan Samani, Petra H. Homoed-van der Kraak, Tim Bezemer, Tim van de Kerkhof, Joyce Vrijenhoek, Evelyn Velema, Ben van Middelaar, Sander Reukema, Robin Reijers, Joëlle van Bennekom, and Bas Nelissen. Lastly, we would like to thank all participants of the Athero-Express Biobank Study; without you these studies would not be possible.

## Supplemental Material

### Supplemental Tables

**Supplemental Table 1:** Athero-Express slide numbers used for the project algorithm design. Some slides were also used for figures. The specific stains used include: CD68 (macrophages), α-smooth muscle actin (a-SMA) (smooth muscle cells), CD66b (neutrophils), CD34 (endothelial cells), glycophorin C (GLYCC) (red blood cells), hematoxylin and eosin (H&E) (nuclei), picrosirius red (SR) (collagen), fibrin (FIBRIN), elastic Van Gieson (EvG). WSI are obtained at 40x magnification, and stored as z-stacked `.TIF` images as produced by the Roche Ventana iScan HT with 0.25 micron/pixel or `.ndpi` images as produced by the Hamamatsu C12000-22 Digital slide scanner with 0.23 micron/pixel.

| Slide number | Stain | Image type | Figure |
| --- | --- | --- | --- |
| AE1299 | CD34 | TIF |  |
| AE1354 | CD34 | TIF |  |
| AE1675 | CD34 | TIF |  |
| AE182 | CD34 | TIF |  |
| AE2086 | CD34 | TIF |  |
| AE2173 | CD34 | TIF |  |
| AE2217 | CD34 | TIF |  |
| AE2622 | CD34 | TIF | Figure 4F |
| AE2637 | CD34 | TIF |  |
| AE2742 | CD34 | TIF |  |
| AE489 | CD34 | TIF |  |
| AE839 | CD34 | TIF |  |
| AE1 | CD68 | TIF |  |
| AE100 | CD68 | TIF | Figure 4E |
| AE100 | CD68 | TIF |  |
| AE1001 | CD68 | TIF |  |
| AE1006 | CD68 | TIF |  |
| AE101 | CD68 | TIF |  |
| AE1010 | CD68 | TIF |  |
| AE1016 | CD68 | TIF |  |
| AE1018 | CD68 | TIF |  |
| AE102 | CD68 | TIF |  |
| AE1299 | EVG | TIF |  |
| AE1354 | EVG | TIF |  |
| AE1675 | EVG | TIF |  |
| AE182 | EVG | TIF |  |
| AE2173 | EVG | TIF |  |
| AE2217 | EVG | TIF |  |
| AE2622 | EVG | TIF |  |
| AE2637 | EVG | TIF | Figure 4H |
| AE2742 | EVG | TIF |  |
| AE3151 | EVG | TIF |  |
| AE489 | EVG | TIF |  |
| AE839 | EVG | TIF |  |
| AE87 | EVG | TIF |  |
| AE1178 | FIBRIN | TIF |  |
| AE1299 | FIBRIN | TIF |  |
| AE146 | FIBRIN | TIF |  |
| AE1675 | FIBRIN | TIF |  |
| AE182 | FIBRIN | TIF |  |
| AE260 | FIBRIN | TIF |  |
| AE489 | FIBRIN | TIF | Figure 4D |
| AE87 | FIBRIN | TIF |  |
| AE99 | FIBRIN | TIF |  |
| AE3606 | GLYCC | ndpi |  |
| AE3756 | GLYCC | ndpi |  |
| AE3884 | GLYCC | ndpi | Figure 4B |
| AE4022 | GLYCC | ndpi |  |
| AE4270 | GLYCC | ndpi |  |
| AE4271 | GLYCC | ndpi |  |
| AE4272 | GLYCC | ndpi |  |
| AE4273 | GLYCC | ndpi |  |
| AE4274 | GLYCC | ndpi |  |
| AE4275 | GLYCC | ndpi |  |
| AE4276 | GLYCC | ndpi |  |
| AE4277 | GLYCC | ndpi |  |
| AE87 | GLYCC | TIF |  |
| AE1007 | H&E | ndpi |  |
| AE1030 | H&E | ndpi |  |
| AE1101 | H&E | ndpi |  |
| AE1189 | H&E | ndpi |  |
| AE1217 | H&E | ndpi |  |
| AE1221 | H&E | ndpi |  |
| AE1222 | H&E | ndpi | Figure 4A |
| AE1227 | H&E | ndpi |  |
| AE1229 | H&E | ndpi |  |
| AE1230 | H&E | ndpi |  |
| AE1237 | H&E | ndpi |  |
| AE1238 | H&E | ndpi |  |
| AE1381 | H&E | ndpi |  |
| AE1382 | H&E | ndpi |  |
| AE1466 | H&E | ndpi |  |
| AE1468 | H&E | ndpi |  |
| AE1473 | H&E | ndpi | Figure 3 |
| AE1490 | H&E | ndpi |  |
| AE1499 | H&E | ndpi |  |
| AE1504 | H&E | ndpi |  |
| AE1299 | a-SMA | TIF |  |
| AE1354 | a-SMA | TIF |  |
| AE1675 | a-SMA | TIF |  |
| AE2086 | a-SMA | TIF |  |
| AE2173 | a-SMA | TIF |  |
| AE2217 | a-SMA | TIF |  |
| AE2622 | a-SMA | TIF | Figure 4G |
| AE2637 | a-SMA | TIF |  |
| AE2742 | a-SMA | TIF |  |
| AE3151 | a-SMA | TIF |  |
| AE489 | a-SMA | ndpi |  |
| AE839 | a-SMA | TIF |  |
| AE1299 | SR | TIF |  |
| AE1354 | SR | TIF | Figure 4C |
| AE182 | SR | TIF |  |
| AE2086 | SR | TIF |  |
| AE2217 | SR | TIF |  |
| AE2622 | SR | TIF |  |
| AE2637 | SR | TIF |  |
| AE2742 | SR | TIF |  |
| AE489 | SR | TIF |  |

**Supplemental Table 2:** Baseline characteristics of study population. Baseline characteristics for the patients included in this study. Age: age in years at the time of inclusion. Note that we report the data for 56 patients. As explained in the Methods section under ‘Patient population’, plaque-material are considered ‘waste biomaterial’ and are allowed to be used without any personal information. In this study we also considered this ‘waste biomaterial’, hence no clinical information, e.g., age, is given here nor when sharing data of the relevant samples (n=3).

| Variable | Description | Overall |
| --- | --- | --- |
| N |  | 56 |
| Age (mean ± s.d.) | | 69.500<br>(9.260) |
| Gender (N, %) | *female* | 33.9 (19) |
|  | *male* | 66.1 (37) |
| Hospital (N, %) | *St. Antonius, Nieuwegein* | 21.4 (12) |
|  | *UMC Utrecht* | 78.6 (44) |
| Artery type (N, %) | *carotid (left & right)* | 85.7 (48) |
|  | *femoral/iliac (left, right or both sides)* | 12.5 (7) |
|  | *other arteries (renal, popliteal, vertebral)* | 1.8 (1) |
| Overall plaque phenotype | *atheromatous* | 21.4 (12) |
|  | *fibroatheromatous* | 19.6 (11) |
|  | *fibrous* | 41.1 (23) |

